# Association of blood pressure with cognitive impairment in elderly hypertensive patients in China: A cross-sectional study

**DOI:** 10.1101/2023.08.15.23294145

**Authors:** Yao Jia, Shige Qi, Peng Yin, Maigeng Zhou, Fusui Ji, Zhihui Wang, Xue Yu

## Abstract

**Aims:** To investigate the prevalence of cognitive impairment among elderly Chinese patients with hypertension and explore the relationship between blood pressure and cognitive impairment. Additionally, the study examined the characteristics of cognitive impairment in hypertensive patients and its impact on blood pressure control.

**Methods:** A cross-sectional study was conducted involving 23,271 subjects. Data collection involved questionnaires, physical examinations, and blood tests. AD8 questionnaire, MMSE questionnaire, and neurologist interview, was used to screen patients for cognitive disabilities. Risk factors and prevalence of cognitive impairment were compared between hypertensive and non-hypertensive groups. The prevalence of cognitive impairment at different blood pressure levels was analyzed within the hypertension group. Differences in MMSE scores across dimensions were compared between hypertensive and non-hypertensive groups.

**Results:** The hypertensive group exhibited additional risk factors for cognitive impairment and a higher prevalence of cognitive impairment. The prevalence of cognitive impairment among hypertensive patients followed a U-shaped curve with increasing blood pressure, with the lowest rate observed among patients with blood pressure in the range of 120-139/80-89 mmHg. Several dimensions of cognitive function were significantly diminished in individuals with hypertension. Patients with cognitive impairment demonstrated lower compliance with hypertension treatment and poorer blood pressure control.

**Conclusion:** Cognitive decline across multiple dimensions is typical among older Chinese patients with hypertension, and cognitive impairment has detrimental effects on blood pressure management. Maintaining blood pressure in the 120-139/80-89 mmHg range appears to be more favorable for preserving cognitive function in hypertensive patients.

## Introduction

The prevalence of mild cognitive impairment (MCI) and dementia among individuals over 60 years old in China is reported to be 15.5% and 6.0%, respectively. Hypertension has been identified as a risk factor associated with cognitive impairment.^1,2^ Previous studies have demonstrated an inverse correlation between blood pressure and the prevalence of cognitive impairment.^3^ Furthermore, strict blood pressure management programs have been proposed to mitigate cardiovascular and cerebrovascular events in the oldest old population.^4^ However, certain scholars have raised concerns regarding the potential exacerbation of cognitive dysfunction through intensive antihypertensive therapy.^5^ To better understand the optimal blood pressure targets for preserving cognitive function in elderly hypertensive patients in China, we conducted a cross-sectional survey. This study aimed to assess the prevalence of cognitive impairment in the elderly hypertensive population, explore the relationship between blood pressure levels and cognitive impairment prevalence, investigate the characteristics of cognitive impairment in hypertensive patients, and examine the impact of cognitive impairment on blood pressure control.

## Methods

### Study population

The study population consisted of participants who were enrolled in the Prevention and Intervention of Neurodegenerative Diseases in the Elderly in China (PINDEC) study, which was initiated in 2015. The primary objective of this study was to investigate the epidemiology of neurodegenerative diseases and identify associated risk factors among individuals aged 60 years and above in China. The investigation areas included six regions: Beijing, Shanghai, Hubei, Sichuan, Yunnan, and Guangxi. To ensure a representative sample, we employed a comprehensive multistage stratified cluster random sampling method which resulting in the selection of 26,164 subjects for inclusion in the study. Previously published report contain detailed information about the study design, sampling methodology, and participant enrollment.^6^ We obtained data from 23,271 participants who had complete blood pressure measurements.

### Data collection

Within the study areas, data collection procedures included questionnaire surveys, physical measurements, and blood sample collection. Standardized questionnaires were used to document detailed demographic information, personal characteristics, and behavioral risk factors such as smoking, drinking, diet, and physical activity. We also obtained data on chronic disease diagnosis and treatment history. The anthropometric parameters of the participants were assessed through physical measurements. Height and weight were measured using an electronic scale, while waist circumference was measured at the horizontal position of the midpoint between the lower margin of the costal arch and the iliac crest of the midaxillary line using a tape measure. Blood pressure and heart rate were measured on the left upper arm using an electronic sphygmomanometer. Before measurement, participants were instructed to rest for 5 minutes in a quiet and suitable indoor environment. Three measurements of blood pressure and heart rate were taken at 1-minute intervals, and the average values were recorded as the absolute blood pressure and heart rate measurements. A three-stage cognitive assessment was administered to all participants, excluding those who self-reported a previous diagnosis of dementia. Participants who scored ≥2 on the 8-item Dementia Screening Questionnaire(AD8)^7^ underwent a Mini-Mental State Examination (MMSE).^8^ Those with an abnormal MMSE score, as defined below, underwent further evaluation by a neurologist to determine whether they met the diagnostic criteria for dementia based on the fourth edition of the Diagnostic and Statistical Manual of Mental Disorders.^9^ Blood samples were collected to measure fasting blood glucose, total cholesterol, total triglyceride, low-density lipoprotein cholesterol, and high-density lipoprotein cholesterol levels. The blood samples were obtained by a trained and validated staff member following a standardized protocol. The collection occurred either at the community health service station where the participants resided or at their households to ensure convenience and comfort.

### Definitions

For this study, advanced age was defined as individuals aged ≥80 years. Body mass index (BMI) was calculated as weight (kg) divided by height squared (m^2^). Adequate exercise was defined as engaging in activities such as walking, playing ball, running, qigong, or farming for at least 10 minutes three to four times a week. Insufficient exercise referred to exercise below this threshold, while sufficient exercise indicated exercise exceeding this threshold. History of smoking was defined as having smoked, and history of drinking was defined as consuming alcoholic beverages more than once per month. Participants self-reporting a diagnosis of hypertension were categorized as the hypertension (HT) group, while those without a hypertension diagnosis were classified as the non-hypertension (non-HT) group. Within the HT group, subjects with a mean blood pressure below 140/90 mmHg were considered “well-controlled,” while those with a mean blood pressure equal to or exceeding these values were classified as “poorly controlled.” Abnormal MMSE scores were defined based on education level: illiterate individuals with scores ≤17 points, those with primary school education with scores ≤20 points, and individuals with junior middle school or above with scores ≤24 points. The MMSE scale was further divided into different dimensions: items 1 to 10 represented spatio-temporal orientation, items 11 to 13 represented immediate memory ability, items 14 to 18 represented attention ability, items 19 to 26 represented language ability, items 27 represented visuo-spatial ability, and items 28 to 30 represented delayed memory ability.^8^ In this study, individuals with AD8 scores ≥2, abnormal MMSE scores, or a diagnosis of dementia were collectively referred to as “cognitive impairment”.

### Statistical methods

This retrospective study employed rigorous statistical methods for data analysis. We used the Wilcoxon rank sum test to compare variables between different groups, which is suitable for analyzing non-parametric data. For continuous data that followed a normal distribution, we utilized analysis of variance (ANOVA). For categorical data, we employed the chi-square test to determine inter-group comparisons. Additionally, the Cochran-Armitage trend test was applied to evaluate trends in categorical data. Prevalence calculations considered sampling weighting, non-response weighting, and post-stratification weighting to derive a final weight for each study participant. All statistical analyses were conducted using SAS 9.4 software. We considered *p*<0.05 to be statistically significant.

## Results

### Risk factors for cognitive impairment in the HT group

Among the total population, 12,838 individuals (55.1%) were identified as hypertensive patients. The HT group had a higher proportion of elderly individuals, those without family pension support, inadequate exercise levels, and lower education levels than the non-HT group. Furthermore, the mean values of BMI and total triglycerides were significantly higher in the HT group compared to the non-HT group. Conversely, the non-HT group displayed a higher proportion of individuals who reported smoking and drinking habits (see Supplemental Table 1 for details).

### Higher prevalence of cognitive impairment in hypertensive patients

In the three-stage cognitive function evaluation, 3,749 individuals had an AD8 score of ≥2, 1,331 individuals had an abnormal MMSE score, and 609 individuals were diagnosed with dementia. The prevalence of AD8 ≥2, MMSE abnormality, and dementia was significantly higher in the HT group compared to the non-HT group (16.99% vs. 15.03%, p<0.001; 6.34% vs. 4.96%, p<0.001; 2.80% vs. 2.39, p<0.05), as illustrated in Figure 1 and Supplemental Table 2. When analyzing the data by age groups, the prevalence of dementia was significantly higher in the HT patients than in the non-HT patients within the 60-69 age group. However, the difference was insignificant in the other age groups, as shown in Figure 2 and Supplemental Table 3.

**Figure 1:**
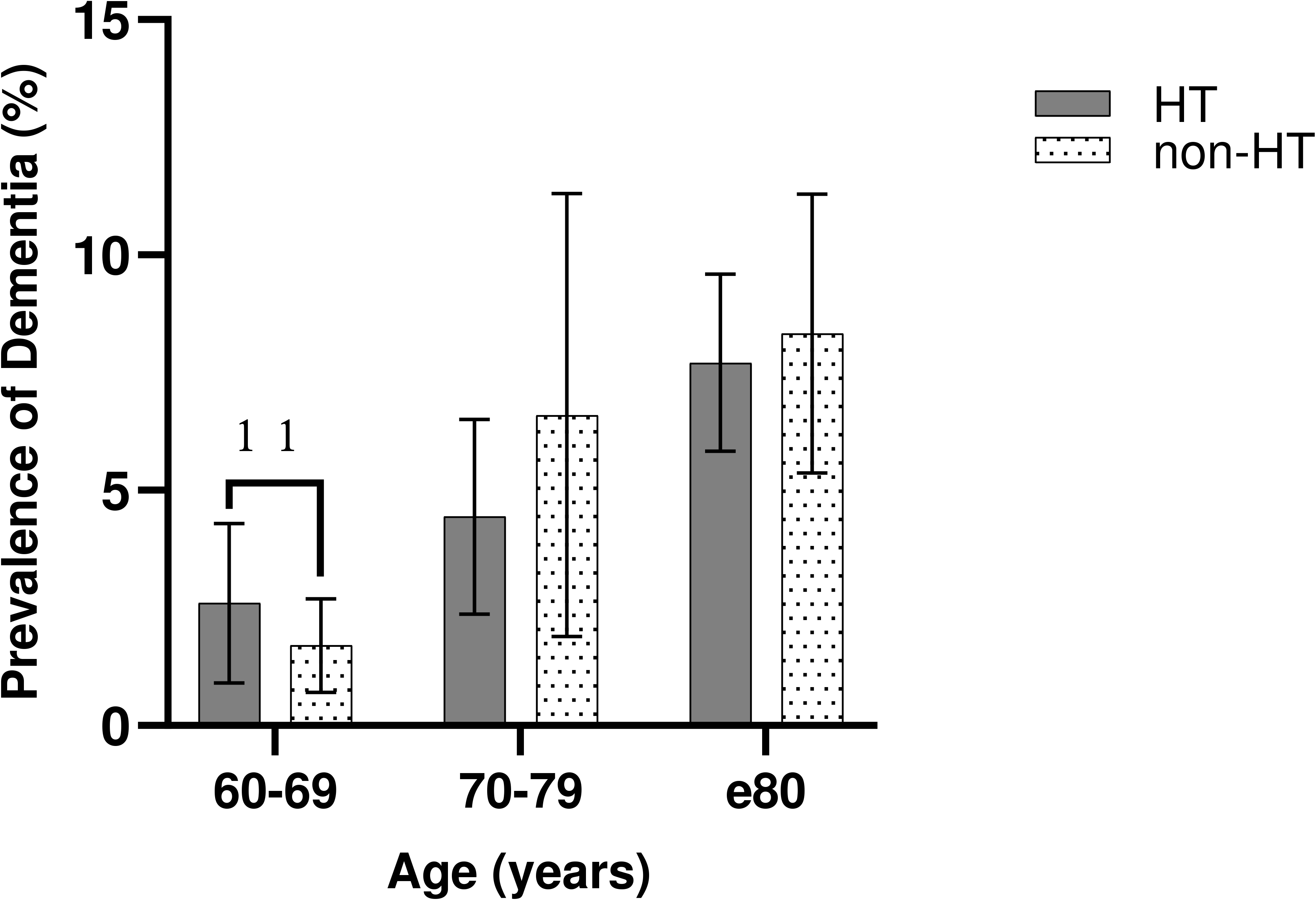
Prevalence of cognitive impairment using various definitions

**Figure 2:**
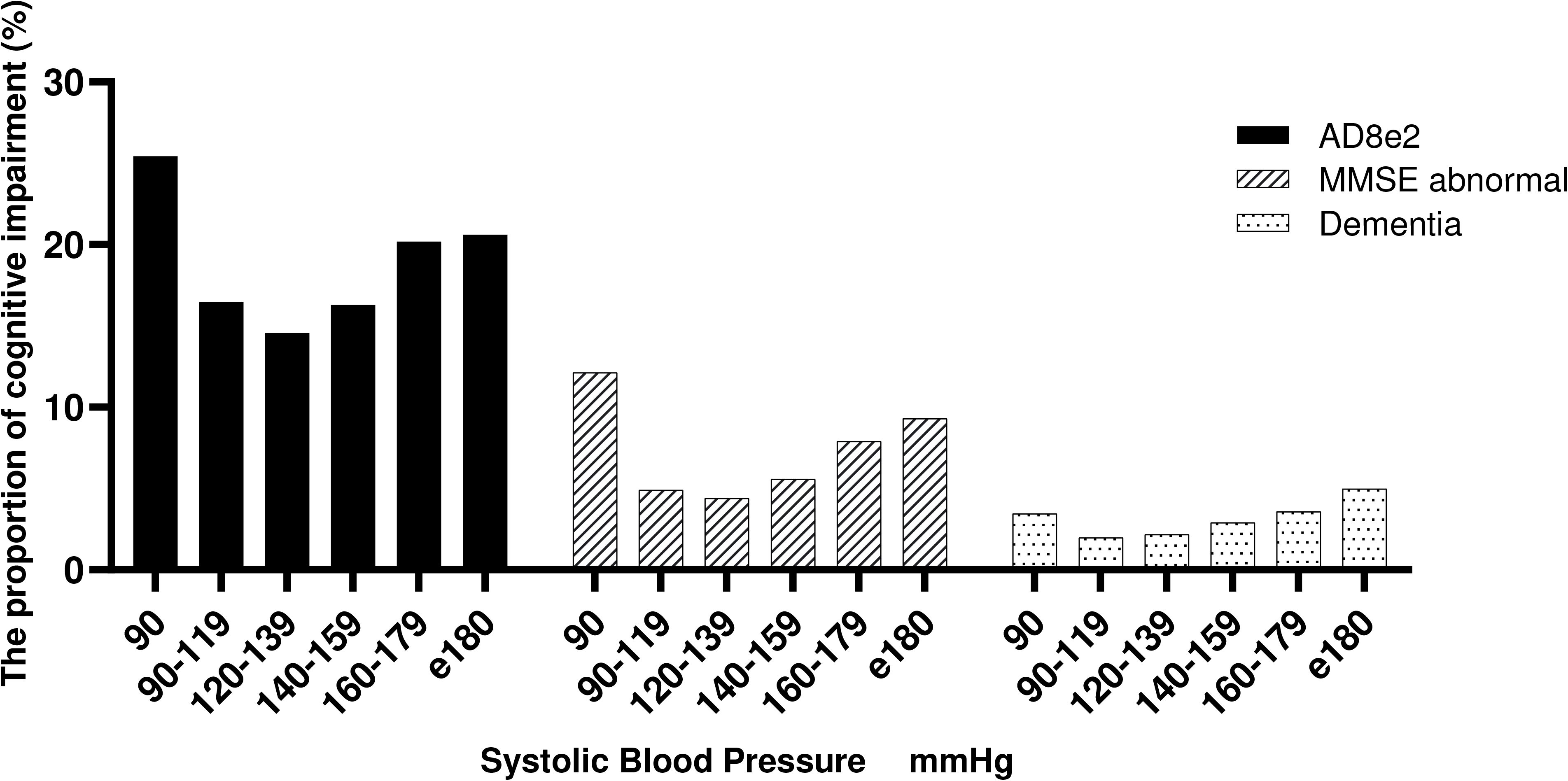
Prevalence of dementia across different age groups

### Distribution of cognitive impairment in the HT group shows a U-shaped pattern with an increase in blood pressure

To investigate the relationship between blood pressure levels and cognitive impairment, hypertensive participants who adhered to their prescribed treatment were classified into six groups based on systolic blood pressure (SBP) in 20 mmHg increments and diastolic blood pressure (DBP) in 10 mmHg increments. The proportion of individuals with AD8 ≥2, abnormal MMSE scores, and dementia in each blood pressure group is presented in Supplemental Tables 4 and 5. Notably, the group with the lowest proportion of cognitive impairment was observed in the blood pressure range of 120-139/80-89 mmHg. As blood pressure levels deviated below or above this range, the proportion of cognitive impairment increased, exhibiting a U-shaped distribution. Figures 3 and 4 show a visual representation of the relationship between cognitive impairment and SBP and DBP level in hypertensive patients.

**Figure 3:**
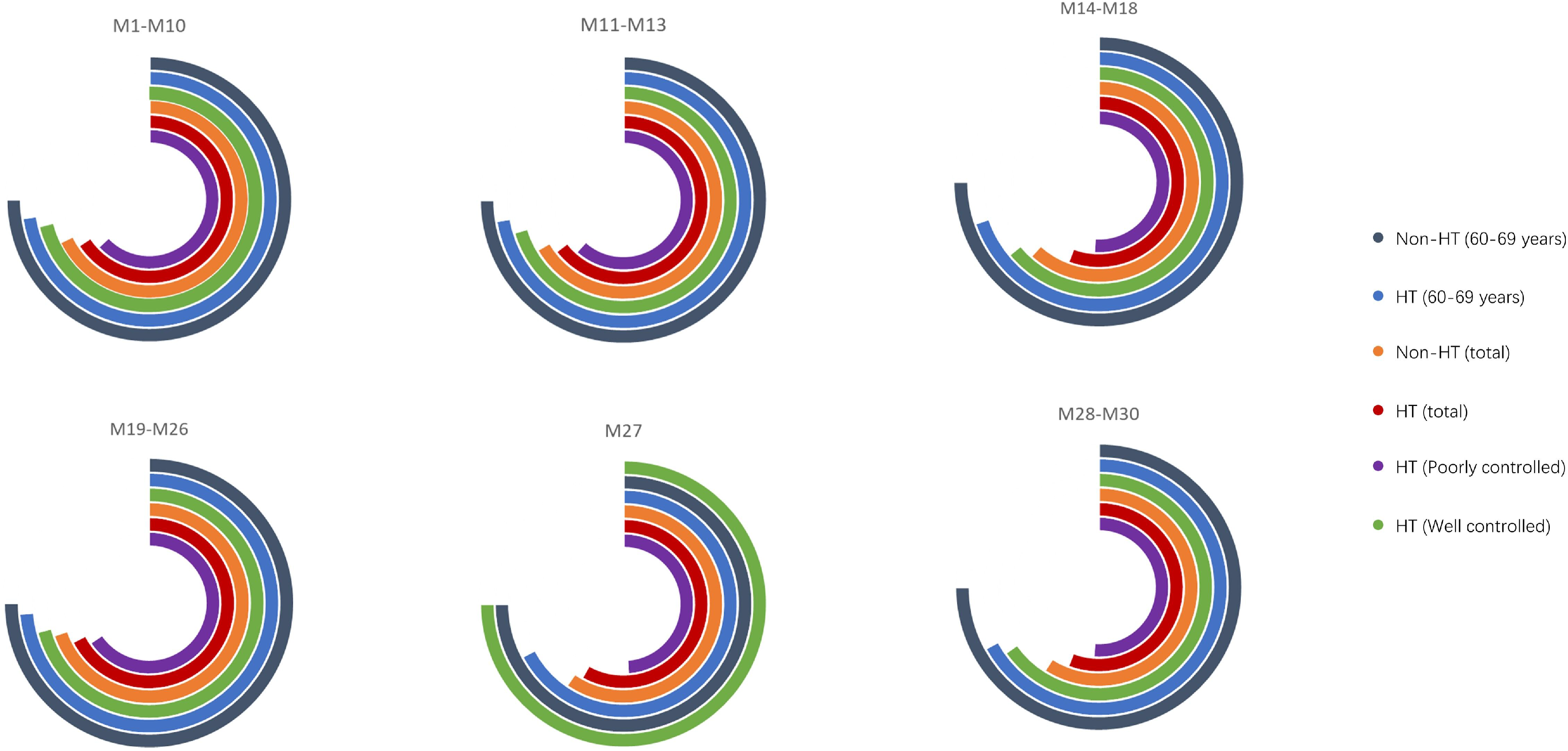
Proportion of cognitive impairment in hypertensive patients based on systolic blood pressure levels

**Figure 4:**
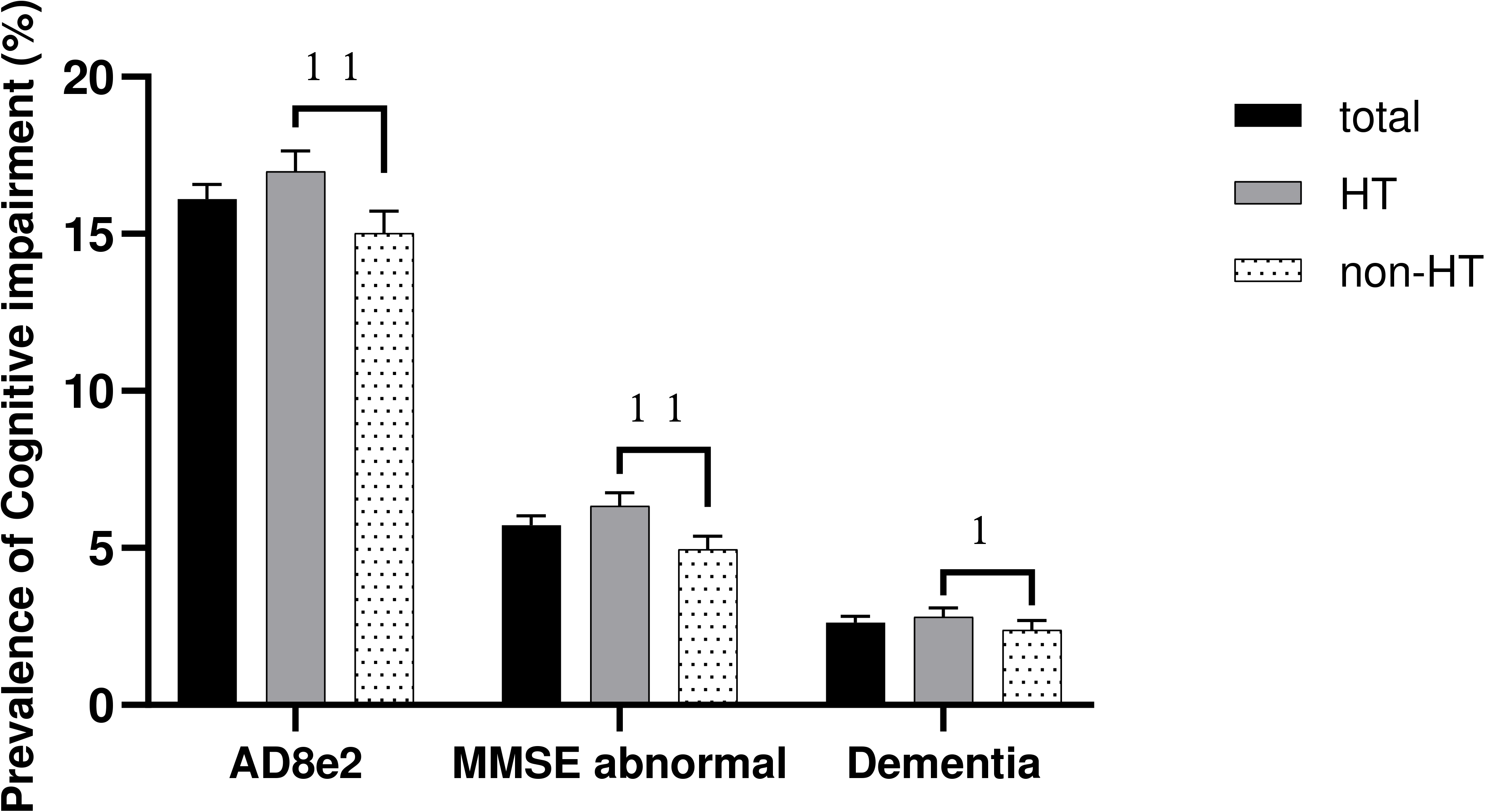
Proportion of cognitive impairment in hypertensive patients based on diastolic blood pressure levels

### Multidimensional cognitive impairment in hypertensive patients

Significant differences were observed in the MMSE scores between the HT and non-HT groups across various cognitive domains, including spatial-temporal orientation, immediate memory, attention, and language ability. Although the 60-69 years old group exhibited higher MMSE scores in all dimensions compared to the overall population, the HT group in this age group still demonstrated lower scores than the non-HT group. Specifically, significant differences were observed in spatial-temporal orientation, immediate and delayed memory, attention, and language ability. In the HT group, cognitive function scores in all domains were significantly decreased in the poorly controlled group, as shown in Figure 5 and summarized in Supplemental Table 6.

**Figure 5:**
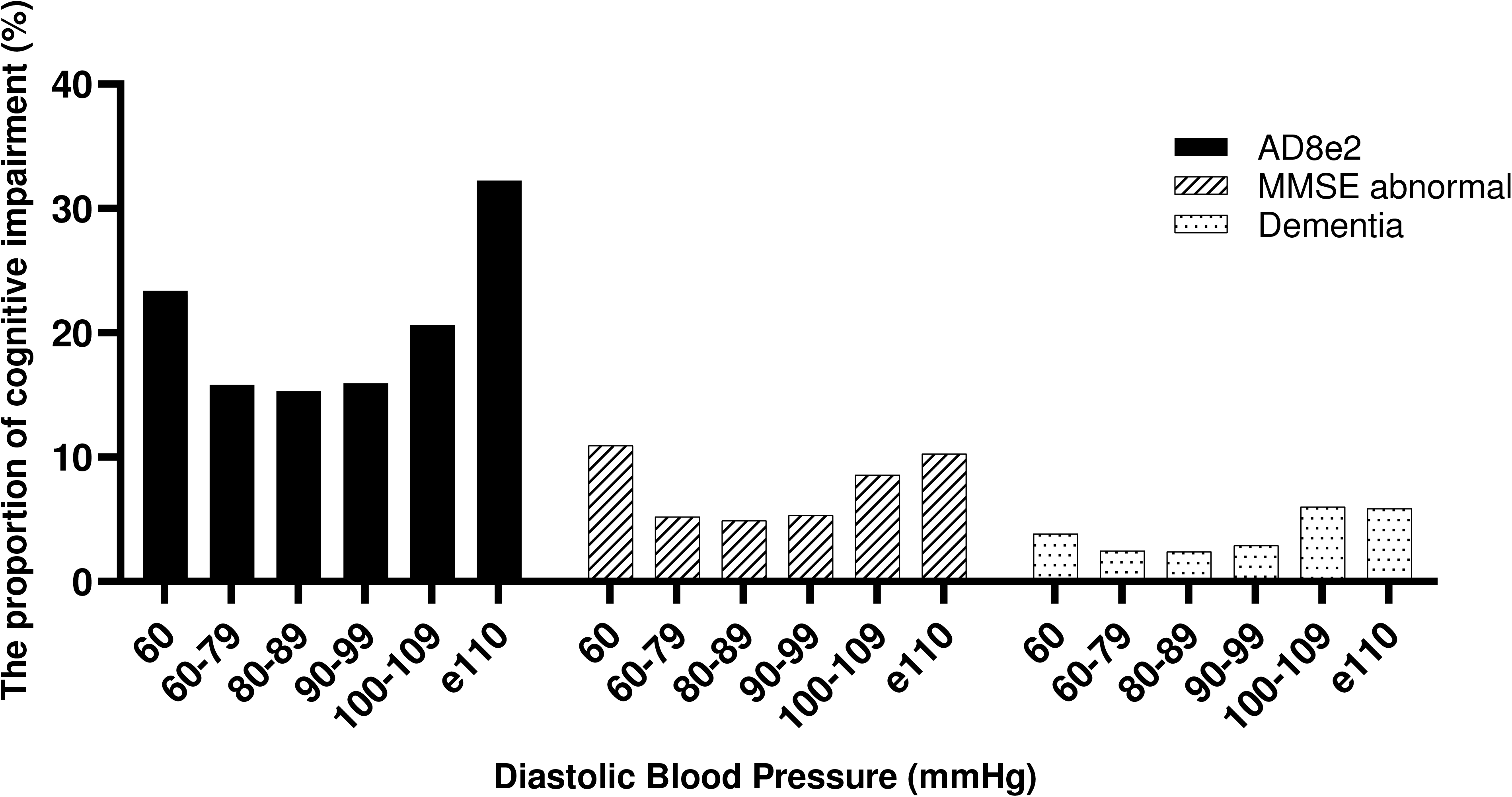
MMSE scores in different cognitive domains among hypertensive and non-hypertensive groups

### Cognitive impairment in hypertensive patients: poor treatment compliance and blood pressure control

Among the HT group, 60.6% of the patients could adhere to their prescribed medication; however, 64.3% of patients had uncontrolled blood pressure. Patients with cognitive impairment exhibited a higher proportion of poor blood pressure control and lower medication adherence rates. Detailed information can be found in Table 1. Additionally, the hypertension group was categorized into four groups based on the quartile of the MMSE total score, and the proportion of poorly controlled blood pressure in each group is presented in Supplemental Table 7. A significant trend test (p < 0.05) revealed that lower MMSE total scores were associated with a higher proportion of poorly controlled blood pressure.

**Table 1.** Blood pressure control and medication adherence of hypertensive patients.

|  | **Total** | **AD8≥2** | **MMSE** | **dementia** |
| --- | --- | --- | --- | --- |
|  |  |  | **abnormal** |  |
| **Poorly BP control** | 8,259 ( 64.3 ) | 1,455 ( 66.7 ) | 584 ( 71.8 )<sup>b</sup> | 253 ( 70.3 )<sup>c</sup> |
|  |  | <sup>a</sup> |  |  |
| **Follow prescription** | 7,785 ( 60.6 ) | 1,269 ( 58.2 ) | 431 ( 52.9 )<sup>b</sup> | 208 ( 58.0 )<sup>c</sup> |
|  |  | <sup>a</sup> |  |  |

Note: <sup>a</sup>compared with AD8 ≤ 1, $p<0.05$ ; <sup>b</sup>compared with MMSE normal,
$p<0.05$ ; <sup>c</sup>compared with individuals without dementia, $p<0.05$

## Discussion

### Prevalence of cognitive impairment in hypertensive patients

Hypertension and cognitive impairment are prevalent among older individuals, and their prevalence tends to increase with age.^2,10–12^ In this study, the proportion of patients with hypertension accounted for 55.1% of the total population, slightly higher than previously reported.^13,14^ We observed that individuals with hypertension are at a higher risk of cognitive impairment due to various associated factors (such as advanced age, high BMI, non-family care, lack of exercise, low education level, and dyslipidemia).^1,15^ These risk factors may contribute to reduced social activities and limited interpersonal communication, which can accelerate the decline in cognitive function. The prevalence of dementia, AD8 score ≥2, and MMSE score abnormality in the HT group were 2.8%, 17.0%, and 6.3%, respectively, all significantly higher than those in the non-HT group. It is worth noting that despite low levels of tobacco and alcohol consumption among the hypertensive population in this study, this may be attributed to the fact that hypertensive patients have received more education on adopting a healthy lifestyle and modified their detrimental habits.

We found a significant difference in the prevalence of dementia between hypertensive and non-hypertensive patients, specifically in the 60-69 age group, whereas no significant difference was observed in higher age groups. This finding may be attributed to the varied etiological classification of dementia in different age groups. Common causes of dementia include Alzheimer’s disease, vascular dementia, dementia with Lewy bodies, and Parkinson’s disease dementia.^12^ Hypertension-associated cognitive dysfunction mechanisms involve cerebral artery intima-media thickening, vessel lumen stenosis, or cerebral blood flow insufficiency due to abnormal blood pressure fluctuations.^16^ Consequently, individuals with hypertension are more susceptible to developing vascular dementia. With advancing age, multiple types of dementia coexist, leading to a reduced disparity in the prevalence of dementia between hypertensive and non-hypertensive patients. In China, almost half of the individuals aged 60-69 are still employed and serve as primary provider for the household. Thus, routine cognitive impairment screening is necessary for hypertensive patients in this age group.

### Blood pressure levels and cognitive impairment

In recent years, it has been acknowledged that stricter blood pressure management can improve cardiovascular and cerebrovascular outcomes in patients with hypertension, including among the elderly.^17,18^ However, concerns have been raised regarding the potential impact of excessively low blood pressure on cerebral perfusion. In this study, we observed a U-shaped relationship between blood pressure and the prevalence of cognitive impairment. The lowest proportion of cognitive impairment was observed when blood pressure ranged between 120-129/80-89 mmHg, and the prevalence increased when blood pressure deviated from this range. These findings are consistent with previous studies.^19,20^ The U-shaped curve suggests that both excessively low and high blood pressure can disrupt cerebral blood flow regulation, leading to inadequate cerebral perfusion, brain tissue ischemia, and hypoxia, ultimately resulting in cognitive impairment.^21–23^ Blood pressure variability also plays a significant role in cognitive function.^24–26^ In this study, among hypertensive patients who did not adhere to medication, we observed a U-shaped trend in the proportion of cognitive impairment with increasing blood pressure. However, this difference was not statistically significant. This lack of significance may be attributed to the substantial blood pressure variability in individuals who did not follow their prescribed medications. Future studies should consider collecting data on blood pressure variability to investigate its impact on cognitive function. Additionally, previous studies have shown that individuals with hypertension in midlife have a higher prevalence of cognitive impairment in older age, whereas no increased prevalence of cognitive impairment was observed in those who developed hypertension later in life.^27^ These findings suggest that the duration of hypertension may be a risk factor for cognitive impairment. Future studies aiming to explore this association could benefit from collecting data on the duration of hypertension.

### Characteristics of cognitive impairment in hypertensive patients

Cognitive decline in various dimensions is highly prevalent among the elderly population. It is important to note that some patients with cognitive impairment may not recognize their symptoms, and healthcare professionals may miss overlook warning signs due to atypical clinical symptoms. The MMSE scale, which assesses cognitive function across spatial-temporal orientation, immediate and delayed memory, attention, language ability, and visuospatial ability was utilized in this study. The findings demonstrate that hypertensive patients had significantly lower total MMSE scores than non-hypertensive patients, particularly in spatial-temporal orientation, immediate memory, attention, and language ability. To mitigate the potential confounding effects of age, we conducted specific analyses on the 60-69 age group, and the results were consistent with previous studies,^28,29^ indicating that cognitive impairment in hypertensive patients within this age range predominantly affected spatial-temporal orientation, memory, and attention. Additionally, previous studies have suggested that subjective memory complaints, assessed through a single question, can be a risk factor for cognitive impairment in hypertensive individuals.^30^ Simplified cognitive impairment screening scales that focus on memory assessment could facilitate a better screening process for cognitive impairment in hypertensive patients.

### Impact of cognitive impairment on blood pressure management

Managing blood pressure in patients with hypertension often faces challenges, resulting in an unsatisfactory control rate.^14,31,32^ Poor treatment compliance is a significant contributing factor to these suboptimal control rates. Several factors, including hidden symptoms in the early stages of the disease (disease factors), the need to take multiple tablets daily (treatment factors), and missed or incorrect medication intake (self-factors), can influence treatment compliance in hypertension. We found that patients with cognitive impairment, as indicated by AD8≥2 and abnormal MMSE scores, exhibited low medication adherence rates and a high prevalence of poorly controlled blood pressure. Furthermore, an inverse relationship was observed between the MMSE score and the proportion of poorly controlled blood pressure. Cognitive impairment can significantly impact treatment compliance and blood pressure control rates in patients with hypertension. Clinicians should remain vigilant regarding cognitive impairment in patients with poorly controlled blood pressure.

### Strengths and limitations of the study

Our study has several strengths that enhance the robustness of our findings. Firstly, the study involved a large sample size, encompassing over 23,000 participants across six provinces in China. This extensive size enhances the representativeness of our conclusions within the population. Secondly, our utilization of a comprehensive three-stage cognitive function assessment method, rather than relying solely on a single questionnaire, contributes to the accuracy of our findings.

However, it is important to acknowledge the limitations of our study. Firstly, this was a cross-sectional study without follow-up data, which restricts our ability to establish causal relationships between hypertension and cognitive impairment. Secondly, the study area did not include northeast China, potentially introducing a selection bias and limiting the generalizability of our findings to the entire Chinese population. Thirdly, blood pressure data was collected only on the survey day, without incorporating 24-hour ambulatory blood pressure monitoring or home blood pressure measurements. As a result, the actual blood pressure status of the participants may not have been fully reflected, which could impact the accuracy of our conclusions.

### Perspective

Considering the increasing prevalence of hypertension and cognitive impairment in our aging society, it is evident that patients with both hypertension and cognitive impairment tend to exhibit poorer treatment compliance and blood pressure control. Thus, it becomes crucial to prioritize the screening and treatment of cognitive impairment in hypertensive patients, as doing so can positively impact blood pressure management outcomes. Moreover, establishing appropriate blood pressure control goals tailored specifically to patients with cognitive impairment can substantially contribute to preserving cognitive function in patients with hypertension. These endeavors hold significant potential for improving the overall health and well-being of patients with hypertension and cognitive impairment in our aging population.

## Abbreviations

PINDEC: Prevention and Intervention of Neurodegenerative Diseases in the Elderly in China
AD8: 8-item Dementia Screening Questionnaire
MMSE: Mini-Mental State Examination
BMI: Body Mass Index
HT: Hypertension
non-HT: non-Hypertension
CI: Cognitive Impairment
ANOVA: Analysis of Variance
CHD: Coronary Heart Disease
DM: Diabetes Mellitus
TG: Total Triglycerides
SBP: Systolic Blood Pressure
DBP: Diastolic Blood Pressure

## Acknowledgements

None

## Authors’ Contributions

Jia Yao: First author. Conceptualization; Data curation; Formal analysis; Writing - original draft

Shige Qi: Co-first Author. Data curation; Formal analysis

Peng Yin: Project administration; Resources; Supervision

Maigeng Zhou: Project administration; Resources; Supervision

Fusui Ji: Project administration; Supervision

Xue Yu: Corresponding author. Conceptualization; Writing - review & editing; Supervision

Zhihui Wang: Co corresponding author. Writing - review & editing; Supervision

## Data availability

All data needed to assess the conclusions in the paper are available within the paper and/or the Supplementary Materials.

